# Antecedent autonomic symptoms predict contemporary autonomic symptom burden and reduced health-related quality of life after spontaneous coronary artery dissection

**DOI:** 10.64898/2026.04.21.26351434

**Authors:** Marie-Claire Seeley, Dang Xuan Anh Tran, Jessica A Marathe, Shiwani Sharma, Gemma Wilson, Suzanne Atkins, Dennis H Lau, Celine Gallagher, Peter J Psaltis

## Abstract

**Introduction:** Spontaneous coronary artery dissection (SCAD) is frequently accompanied by persistent symptoms of unknown pathogenesis after the index event. Autonomic dysfunction is a plausible mechanism for these but has not been systematically characterized. We quantified antecedent and contemporary autonomic symptoms in survivors of SCAD and examined their associations with cardiac and extra-cardiac symptoms and health-related quality of life.

**Methods:** This cross-sectional study recruited 227 volunteers from multiple countries with a self-reported history of SCAD. Participants completed validated patient-reported measures, including the Composite Autonomic Symptom Score-31 (COMPASS-31), Anxiety Sensitivity Index-3 (ASI–3), and EuroQol-5 Dimension-5L (EQ-5D-5L). They also completed an internally derived retrospective autonomic predisposition score assessing symptoms during adolescence and early adulthood.

**Results:** Participants were predominantly female (97.8%), median age 53 (47–58) years, and were surveyed a median of 3 (1–5) years after their index SCAD event. 21.6% reported SCAD recurrence. Moderate autonomic symptom burden (COMPASS-31 ≥20) was present in 56.4% and severe burden (≥40) in 16.3%. History of antecedent autonomic symptoms was the strongest independent predictor of contemporary autonomic symptom burden after adjustment for demographic and clinical covariates (*β*=0.514; *P* <0.001). Greater autonomic symptom burden independently predicted lower EQ-5D health utility (*β*=−0.150; *P*=0.029) and was associated with the ASI-3 ‘*physical concerns*’ (*β*=0.232; *P* <0.001), but not ‘*social concerns*’ domain. Autonomic symptoms were not associated with SCAD recurrence.

**Conclusion:** Symptoms of autonomic dysregulation are common in survivors of SCAD and are associated with reduced quality of life. Their association with antecedent dysautonomic features during adolescence and early adulthood suggests a longstanding predisposition, the significance of which warrants further evaluation.

**Clinical Perspective:** *What Is New?:*

- Self-reported antecedent and current autonomic symptoms are common in survivors of spontaneous coronary artery dissection and are associated with poorer health-related quality of life, greater fatigue, and greater psychological distress.

*What Are the Clinical Implications?:*

- Autonomic symptoms warrant clinical recognition in patients with prior spontaneous coronary artery dissection, not only as a post-event complaint but also as a potential marker of pre-existing autonomic vulnerability that may influence recovery experience.
- Greater awareness of autonomic symptom burden may support more personalized follow-up, patient counseling, and rehabilitation planning to help patients return more safely and confidently to daily activities, work, and family life.

**Graphical Abstract:** 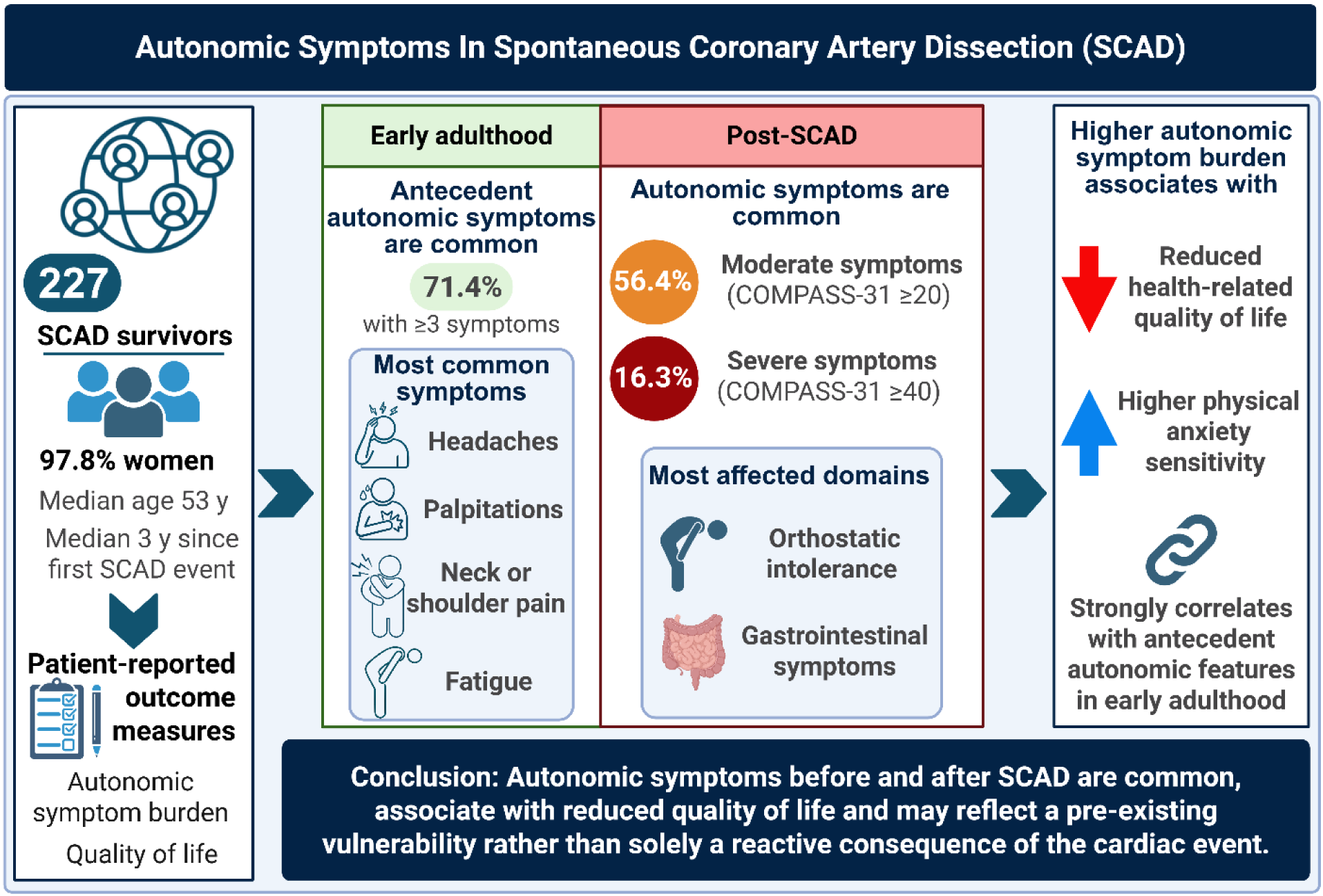

## Introduction

Spontaneous coronary artery dissection (SCAD) occurs when a non-atherosclerotic, non-iatrogenic, non-traumatic intramural hematoma develops with or without intimal tear, causing separation of the coronary arterial wall. This leads to compression of the true lumen of the vessel, compromising myocardial blood flow and resulting in myocardial ischemia and/or infarction.^1^ Contemporary registries report that SCAD accounts for 0.2 to 4% of all acute coronary syndromes and around 25% of acute myocardial infarction (MI) in women under 60 years of age, often in the absence of traditional cardiovascular risk factors.^2–4^ Although the pathogenesis of SCAD remains uncertain, several lines of evidence suggest a vulnerability of the connective tissue within the arterial wall, including a strong association with fibromuscular dysplasia (FMD).^5^ Physical, emotional and physiological stressors associated with sympathetic nervous system activation, such as intense exertion, acute emotional stress, and hormonal fluctuations have been frequently reported as precipitating factors.^3, 6^ Individuals with SCAD often go on to experience a high burden of debilitating cardiac and extra-cardiac symptoms, in addition to psychological distress, beyond their acute event.^7–9^ Qualitative and quantitative research has documented anxiety, depression, post-traumatic stress disorder, uncertainty about recovery, and impaired quality of life that may persist for months to years.^10^

The autonomic nervous system (ANS) is instrumental in vasomotor regulation and cardiovascular homeostasis via its sympathetic and parasympathetic branches and plays a key role in the etiology and progression of cardiovascular disease.^11^ Fluctuations in stressors affecting ANS control of vascular tone can manifest in a myriad of symptoms, including orthostatic intolerance, flushing, palpitations, migraines, and gastrointestinal dysfunction.^11, 12^ Notably, some of these symptoms are highly prevalent in patients with SCAD, hinting that shared neurovascular pathways may contribute to these presentations.^7, 13^ However, until now, dedicated characterization of ANS dysfunction in SCAD has been limited. To address this knowledge gap, we conducted a cross-sectional study quantifying antecedent and current autonomic symptoms in SCAD survivors and examined their associations with cardiac and extra-cardiac manifestations, psychological distress, fatigue, and health-related quality of life.

## Methods

### Data availability

Deidentified participant data will be made available to qualified researchers on reasonable request to the corresponding author, subject to institutional ethics approval, data governance requirements, and participant consent restrictions.

### Study design and participants

Adults aged ≥18 years with a self-reported, clinically confirmed diagnosis of SCAD on invasive coronary angiography or coronary computed tomography angiography were eligible. Participants were recruited through ethically approved advertising via consumer organizations, clinician networks, and online patient communities. A separate cohort of adults with clinician-diagnosed postural orthostatic tachycardia syndrome (POTS) was recruited under the same ethics approval using identical procedures. This cohort was included only for construct validation of the historical autonomic predisposition measure and excluded from primary SCAD analyses. The study complied with the Declaration of Helsinki and the National Statement on Ethical Conduct in Human Research (2023). Ethics approval was granted by the Adelaide University Human Research Ethics Committee (H-2025-066), and all participants provided electronic informed consent.

### Data collection

Data were collected between 21 May 2025 and 31 December 2025 using a secure, web-based platform (Research Electronic Data Capture [REDCap]).^14^ Participants completed self-report questionnaires to characterize autonomic symptom burden and explore its relationship to other commonly reported symptoms in patients with a history of SCAD, including migraine, anxiety-related symptoms, fatigue, and reduced health-related quality of life.

### Diagnostic experience questionnaire

Diagnostic experiences related to SCAD were assessed using a structured, study-specific questionnaire capturing event history, reproductive status at the index event, perceived physical and emotional stressors in the six months preceding the event, diagnostic modality, and delay to diagnosis. Additional items assessed emergency department presentations before diagnosis. The questionnaire was designed to characterize patient-reported diagnostic pathways and health care utilization not routinely captured in clinical registries.

### Patient-reported outcome measures

Autonomic symptom burden was assessed using the Composite Autonomic Symptom Score-31 (COMPASS-31), a validated 31-item instrument capturing symptoms across six domains: orthostatic intolerance, vasomotor, secretomotor, gastrointestinal, bladder, and pupillomotor symptoms.^15^ Domain scores were weighted and summed to generate a total score ranging from 0 to 100, with higher scores indicating greater autonomic symptom burden.^15^ A COMPASS-31 ≥20 was used to define moderate autonomic symptom burden and ≥40 to define clinically severe burden, as previously described.^16, 17^

Antecedent autonomic features were assessed using a 10-item dichotomous questionnaire developed for this study to capture symptoms during adolescence and early adulthood consistent with systemic autonomic dysfunction. Items were informed by recognized clinical phenotypes of autonomic disorders and expert clinical experience.^18^ Affirmative responses were summed to generate a Composite Historical Autonomic Predisposition Score (CHAPS) (range, 0–10), with higher scores indicating greater historical autonomic symptom burden. Internal consistency was assessed using Cronbach’s alpha, and construct validity was evaluated by correlations with current autonomic symptom burden (COMPASS-31) and inclusion in multivariable regression models (**Supplementary Appendix 1**).

Psychological symptoms were assessed using the Anxiety Sensitivity Index–3 (ASI–3) and the Center for Epidemiologic Studies Depression Scale (CES-D). The ASI–3 measures fear of anxiety-related somatic sensations across physical, cognitive, and social domains, including overlap with autonomic symptom perception.^19^ The CES-D is a 20-item measure of depressive symptoms over the previous week (score range, 0–60); positively worded items were reverse scored. Consistent with prior studies in medically ill populations, scores ≥20 indicated clinically significant depressive symptom burden.^20^

Fatigue severity was measured using the Fatigue Severity Scale (FSS), a 9-item instrument assessing the functional impact of fatigue, with total scores calculated as the mean of all items (range 1–7).^21^ Generalized joint hypermobility was assessed using the validated 5-point hypermobility questionnaire described by Hakim and Grahame.^22^ The questionnaire consists of five “yes/no” items evaluating current and historical features of joint laxity. A score ≥2 was used to define generalized joint hypermobility, consistent with established validation studies demonstrating good sensitivity and specificity relative to the Beighton score.^23^

Health-related quality of life was assessed using the EuroQol 5-Dimension (EQ-5D). Responses across five domains were converted to a utility index in which 1 represents full health, 0 death, and values <0 health states considered worse than death.^24, 25^ Utility scores were calculated using the Australian (Norman) EQ-5D-5L value set, selected *a priori* because most participants resided in Australia or other high-income countries with comparable health system contexts.^24^ Participants also completed the EQ visual analogue scale (EQ-VAS), rating overall health from 0 to 100 as a complementary global measure of perceived health status.^25^

### Statistical analysis

Analyses were performed using IBM SPSS Statistics, version 29.0 (IBM Corp., Armonk, NY). Only fully completed surveys were included; no imputation was performed. Continuous variables were assessed for normality and are presented as mean ± SD or median (IQR), as appropriate. Group comparisons used independent samples t-tests or Mann-Whitney U tests. Categorical variables are presented as frequencies and percentages and were compared using chi-square tests.

Associations between autonomic symptom burden and health-related quality of life were assessed using Spearman correlations and linear regression, with standardized coefficients (β) and R² reported. Associations between historical autonomic predisposition and current autonomic symptom burden were assessed using Spearman correlations with COMPASS-31 total and domain scores. Internal consistency was evaluated using Cronbach α. Known-groups validity was examined by comparing historical autonomic predisposition scores between SCAD and POTS cohorts using the Mann-Whitney U test, with effect size calculated as r = Z/√N.

Independent predictors of autonomic symptom burden and EQ-5D-5L utility were examined using hierarchical linear regression with prespecified blocks. For COMPASS-31, blocks included demographics (age, body mass index [BMI], smoking), SCAD-related variables (number of events, years since index event), and biological phenotype variables (historical autonomic predisposition score, irritable bowel syndrome, hypotension). For EQ-5D-5L utility, blocks included demographics, SCAD-specific factors, biological/comorbid variables, COMPASS-31, and CES-D non-somatic score. Associations between autonomic symptom burden and anxiety sensitivity were assessed using Spearman correlations and multivariable linear regression adjusted for depressive symptoms, with a sensitivity analysis of nonphysical ASI domains. Standardized coefficients (β) adjusted R² and change in R² are reported. Prespecified blocks were based on clinical plausibility and prior literature.^26^ Variance inflation factors and tolerance statistics showed no evidence of multicollinearity. All tests were two-tailed, with *P* <0.05 considered statistically significant.

## Results

### Participant characteristics

255 survey responses were received, of which 227 (89%) were fully completed and included in the final analysis. The cohort was predominantly female (97.8%) and of White ethnicity (94.3%), with median age of 53 years (IQR 47–58) (**Table 1**). Participants were mostly resident in Australasia (44.5%), North America (29.1%) and Europe (26.0%), with minimal representation from other regions (0.4%). Fewer than half were in full-time employment or education at the time of survey completion (41.0%), and 7.0% reported being unable to work due to disability. Social isolation was also captured, with 9.7% indicating less than monthly social contact.

**Table 1.**
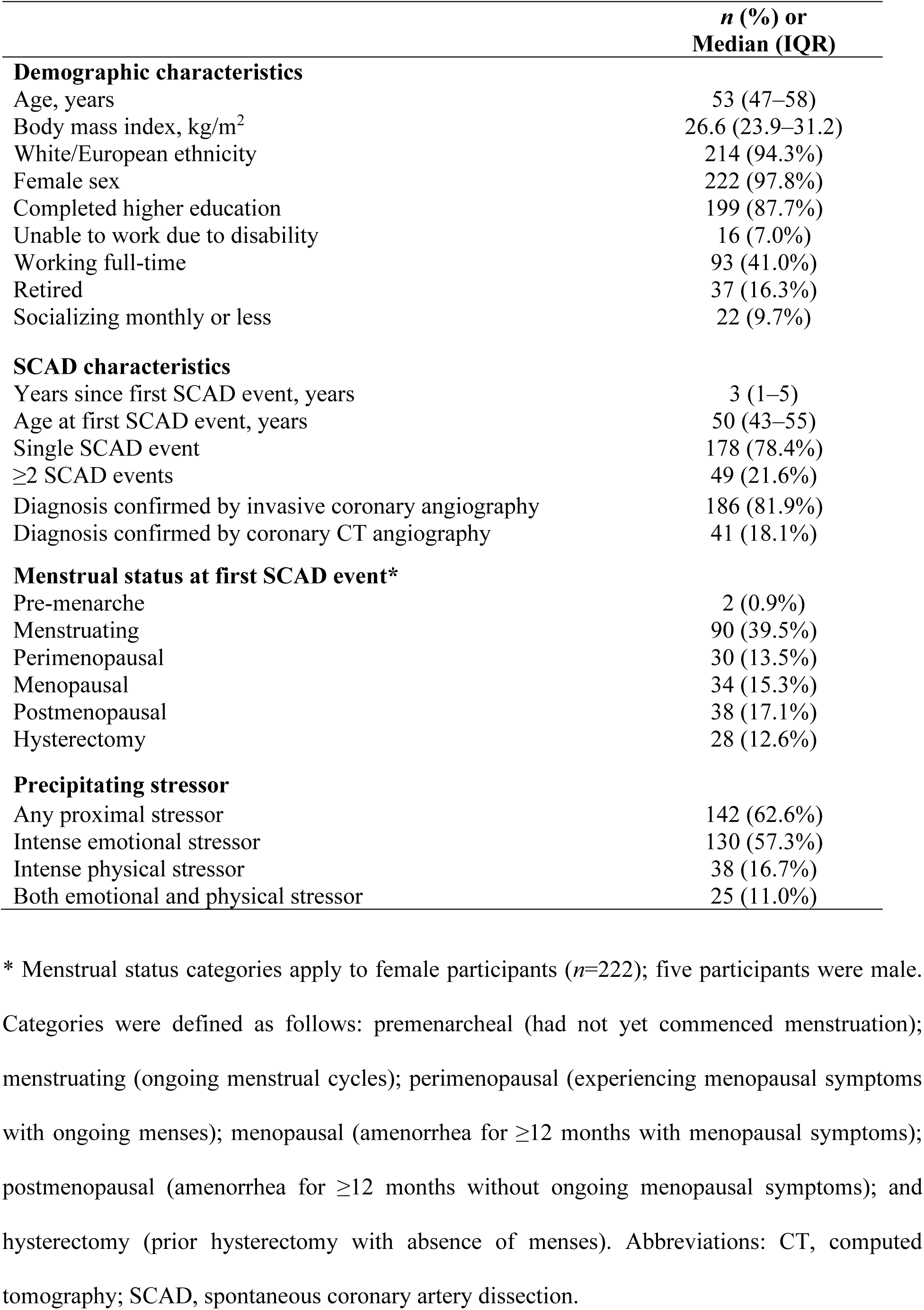
Participant characteristics (*n*=227)

|  | ***n* (%) or**<br>**Median (IQR)** |
| --- | --- |
| **Demographic characteristics** |  |
| Age, years | 53 (47–58) |
| Body mass index, kg/m<sup>2</sup> | 26.6 (23.9–31.2) |
| White/European ethnicity | 214 (94.3%) |
| Female sex | 222 (97.8%) |
| Completed higher education | 199 (87.7%) |
| Unable to work due to disability | 16 (7.0%) |
| Working full-time | 93 (41.0%) |
| Retired | 37 (16.3%) |
| Socializing monthly or less | 22 (9.7%) |
| **SCAD characteristics** |  |
| Years since first SCAD event, years | 3 (1–5) |
| Age at first SCAD event, years | 50 (43–55) |
| Single SCAD event | 178 (78.4%) |
| ≥2 SCAD events | 49 (21.6%) |
| Diagnosis confirmed by invasive coronary angiography | 186 (81.9%) |
| Diagnosis confirmed by coronary CT angiography | 41 (18.1%) |
| **Menstrual status at first SCAD event\*** |  |
| Pre-menarche | 2 (0.9%) |
| Menstruating | 90 (39.5%) |
| Perimenopausal | 30 (13.5%) |
| Menopausal | 34 (15.3%) |
| Postmenopausal | 38 (17.1%) |
| Hysterectomy | 28 (12.6%) |
| **Precipitating stressor** |  |
| Any proximal stressor | 142 (62.6%) |
| Intense emotional stressor | 130 (57.3%) |
| Intense physical stressor | 38 (16.7%) |
| Both emotional and physical stressor | 25 (11.0%) |

\* Menstrual status categories apply to female participants (n=222); five participants were male.
Categories were defined as follows: premenarcheal (had not yet commenced menstruation); menstruating (ongoing menstrual cycles); perimenopausal (experiencing menopausal symptoms with ongoing menses); menopausal (amenorrhea for ≥12 months with menopausal symptoms); postmenopausal (amenorrhea for ≥12 months without ongoing menopausal symptoms); and hysterectomy (prior hysterectomy with absence of menses). Abbreviations: CT, computed tomography; SCAD, spontaneous coronary artery dissection.

Traditional cardiovascular risk factors were relatively infrequent. Median BMI was 26.6 kg/m² (IQR 23.9–31.2). Current smoking was uncommon (2.2%), although 30.0% reported previous smoking. Alcohol use was reported by 60.8% of participants, most commonly at monthly or less (34.1% of drinkers) or 2–4 times per month (30.4%), with 35.5% reporting consumption ≥2 times per week. A diagnosis of hypertension was present in 26.0% and dyslipidemia in 17.6% of participants.

### Characteristics of index SCAD event

Median time between the index SCAD event and survey completion was 3 years (1–5). Most participants (81.9%) reported diagnosis by invasive coronary angiography. The majority (78.4%) experienced a single SCAD event, with 21.6% reporting recurrence. Median age at first SCAD was 50 years (43–55). At the time of the index event, 40.4% of women were premenopausal, 13.5% perimenopausal and 32.4% menopausal or postmenopausal; 12.6% had undergone hysterectomy (**Table 1**). A diagnostic delay exceeding 24 hours was reported by 41.0%, and 62.1% presented to an emergency department at least once for chest pain prior to receiving a SCAD diagnosis.

62.6% of participants recalled a perceived precipitating trigger within the six months preceding the index SCAD event. Intense emotional stress was more frequently reported than intense physical stress (57.3% vs 16.7%). In those identifying a specific emotional trigger (*n*=124; 54.6% of the cohort), the most frequently reported precipitants were work-related stress or interpersonal conflict (31.5%) and relationship breakdown (30.6%), followed by bereavement (17.7%) and family illness (11.3%). Among individuals reporting a physical trigger with further specification available (*n*=35), intense weightlifting (31.4%) and vigorous cardiovascular exertion (28.6%) were most common. Smaller proportions reported pregnancy-related events (14.3%), other intense physical events (20.0%), or severe vomiting (5.7%).

In analyses comparing participants with a single event (*n*=178) to those with ≥2 events (*n*=49), recurrent SCAD was associated with younger age at first event (47 [39–54] vs 50 [43–55] years, *P*=0.020), longer time since index event (5 [2–11.5] vs 2 [1–4] years, *P* <0.001), and higher migraine prevalence (*P*=0.020) (**Supplementary Table 1**). Autonomic symptom burden (COMPASS-31 total and sub-domains), CHAPS, and anxiety sensitivity were not associated with recurrence (all *P* >0.05).

### Autonomic symptom burden

Autonomic symptom burden in the 12 months preceding survey completion was substantial (mean COMPASS-31 24.3±15.5). Based on COMPASS-31 thresholds, 56.4% of participants had scores ≥20, indicating at least moderate autonomic symptom burden, and 16.3% had scores ≥40, consistent with severe symptoms. Cardiovascular (orthostatic intolerance: 10.2±10.4) and gastrointestinal domains (6.8±4.4) contributed most to overall burden, followed by secretomotor symptoms (3.8±3.5) (**Table 2**).

**Table 2.** Autonomic and quality of life characteristics of the cohort (*n*=227)

| **Variable** | **Mean ± SD or Median (IQR)**<br>**or *n* (%)** |
| --- | --- |
| **Autonomic symptom burden** |  |
| COMPASS-31 total score (/100) | 24.3 ± 15.5 |
| Orthostatic intolerance (/40) | 10.2 ± 10.4 |
| Vasomotor (/5) | 0.5 ± 1.1 |
| Secretomotor (/15) | 3.8 ± 3.5 |
| Gastrointestinal (/25) | 6.8 ± 4.4 |
| Bladder (/10) | 1.1 ± 1.4 |
| Pupillomotor (/5) | 1.8 ± 1.1 |
| COMPASS-31 ≥ 20 | 128 (56.4%) |
| COMPASS-31 ≥ 40 | 37 (16.3%) |
| **Health-related quality of life and fatigue** |  |
| EQ-VAS | 71.0 (50.0–80.0) |
| EQ-5D index | 0.924 (0.848–0.956) |
| Fatigue Severity Scale | 39.0 ± 15.5 |
| Severe fatigue (FSS ≥ 36) | 128 (56.4%) |

Severe fatigue was defined as a Fatigue Severity Scale score ≥ 36, consistent with validated thresholds. Abbreviations: COMPASS-31, Composite Autonomic Symptom Score-31; EQ-VAS, EuroQol Visual Analogue Scale; EQ-5D index, EuroQol five-dimension index utility score; FSS, Fatigue Severity Scale; IQR, interquartile range; SD, standard deviation.

### Historical autonomic predisposition

The CHAPS, developed to capture antecedent systemic autonomic signs and symptoms, demonstrated strong known-groups validity, with significantly higher scores in the POTS cohort compared with the SCAD cohort (median 9 [IQR 8–10] vs 4 [IQR 2–6]; *P* <0.001), reflecting a large effect size (*r*=0.71). In the SCAD cohort, antecedent autonomic symptoms were common. 91.6% of participants reported ≥1 symptom and 71.4% reported ≥3 symptoms during adolescence or young adulthood. The mean CHAPS in this cohort was 4.2±2.7. The most frequently reported symptoms were headaches (59.9%), palpitations (51.1%), neck or shoulder pain (48.9%), fatigue (48.5%) , gastrointestinal symptoms (47.6%), exercise-associated flushing (45.4%) and heat intolerance (42.3%). Prior orthostatic dizziness was reported by 37.4% of participants, whereas dependent acrocyanosis was less frequent (17.6%). Internal consistency was acceptable (*Cronbach’s α*=0.76).

CHAPS total score moderately correlated with the COMPASS-31 total score (*ρ*=0.559, 95%CI=0.459–0.644, *P* <0.001). Domain-specific correlations were observed for orthostatic intolerance (*ρ*=0.416), gastrointestinal (*ρ*=0.482), pupillomotor (*ρ*=0.391), and secretomotor (*ρ*=0.332) domains (all *P* <0.001), with smaller but significant associations for vasomotor (*ρ*=0.271) and bladder (*ρ*=0.271) domains (both *P* <0.001) **(Table 3).**

**Table 3.** Associations between historical autonomic symptoms and current autonomic symptom burden (n=227)

| **Outcome** | **Spearman $\rho$** | **95% *CI*** | ***P* value** |
| --- | --- | --- | --- |
| COMPASS-31 total | 0.559 | 0.459–0.644 | **<0.001** |
| Orthostatic intolerance | 0.416 | 0.298–0.521 | **<0.001** |
| Vasomotor | 0.271 | 0.142–0.390 | **<0.001** |
| Secretomotor | 0.332 | 0.207–0.446 | **<0.001** |
| Gastrointestinal | 0.482 | 0.372–0.579 | **<0.001** |
| Bladder | 0.271 | 0.142–0.391 | **<0.001** |
| Pupillomotor | 0.391 | 0.271–0.499 | **<0.001** |

Values represent Spearman rank correlation coefficients ( $\rho$ ) with 95% confidence intervals.
Historical autonomic symptoms were assessed using the Composite Historical Autonomic
Predisposition Score (CHAPS). Current autonomic symptom burden was measured using the
Composite Autonomic Symptom Score-31 (COMPASS-31). All tests were two-sided.

### Predictors of autonomic symptom burden

In hierarchical linear regression analysis, demographic variables explained 10.1% of the variance in COMPASS-31 score (*adjusted R*²=0.089, *P* <0.001) (**Supplementary Table 2**). Younger age (*β*=-0.248, *P* <0.001) and smoking (*β*=0.183, *P*=0.005) were independently associated with greater autonomic symptom burden, whereas BMI did not meet statistical significance (*P*=0.051). Addition of SCAD-related variables did not significantly improve model fit (Δ*R*²=0.014, *P*=0.169), whereas inclusion of biological phenotype variables did (Δ*R*²=0.267, *P* <0.001), with the final model explaining 36.0% of variance in COMPASS-31 score (*adjusted R*²=0.360). Historical autonomic predisposition (CHAPS) demonstrated the strongest independent association with COMPASS-31 score (*β*=0.514, *P* <0.001). Diagnosis of hypotension (*β*=0.126, *P*=0.022) and smoking (*β*=0.144, *P*=0.009) were also independently associated with greater autonomic symptom burden.

### Frequency of comorbidities

Practitioner-diagnosed comorbidities, namely migraine (38.8%), anxiety (34.4%), and depression (33.0%), were common in the SCAD cohort (**Table 4)**. Generalized joint hypermobility (score ≥2) was present in 33.9% of participants. Self-reported prevalence of FMD was 21.6% overall and varied by geographic region (χ²=12.65, *P*=0.005), with a higher rate among North American participants (36.4%) compared with Australasia (13.9%) and Europe (18.6%).

**Table 4.** Comorbid conditions and cardiovascular risk factors according to autonomic symptom burden.

| Condition | Whole cohort | COMPASS-31<br><20 (n=99) | COMPASS-31<br>≥20 (n=128) | *P* value |
| --- | --- | --- | --- | --- |
| **Autonomic-associated conditions** |  |  |  |  |
| Migraine | 88 (38.8%) | 31 (31.3%) | 57 (44.5%) | **0.043** |
| Irritable bowel syndrome | 25 (11.0%) | 6 (6.1%) | 19 (14.8%) | **0.036** |
| Hypotension | 15 (6.6%) | 2 (2.0%) | 13 (10.2%) | **0.014** |
| Asthma | 36 (15.9%) | 9 (9.1%) | 27 (21.1%) | **0.014** |
| Tinnitus | 30 (13.2%) | 7 (7.1%) | 23 (18.0%) | **0.016** |
| Generalized joint hypermobility | 77 (33.9%) | 26 (26.3%) | 51 (39.8%) | **0.032** |
| **Affective diagnoses** |  |  |  |  |
| Anxiety | 78 (34.4%) | 27 (27.3%) | 51 (39.8%) | **0.048** |
| Depression | 75 (33.0%) | 25 (25.3%) | 50 (39.1%) | **0.028** |
| **Vascular arteriopathy** |  |  |  |  |
| Fibromuscular dysplasia | 49 (21.6%) | 20 (20.2%) | 29 (22.7%) | 0.656 |
| **Cardiovascular risk factors** |  |  |  |  |
| Dyslipidemia | 40 (17.6%) | 21 (21.2%) | 19 (14.8%) | 0.212 |
| Hypertension | 59 (26.0%) | 27 (27.3%) | 32 (25.0%) | 0.699 |
| Any alcohol use | 138 (60.8%) | 64 (64.6%) | 74 (57.8%) | 0.296 |
| Current or former smoker | 73 (32.2%) | 23 (23.2%) | 50 (39.1%) | **0.011** |
| BMI ≥30 kg/m<sup>2</sup> | 75 (33.0%) | 30 (30.3%) | 45 (35.2%) | 0.441 |

Values are presented as *n* (%). Autonomic symptom burden was categorized using COMPASS-
31 <20 vs ≥20. *P* values are derived from two-sided Pearson chi-square tests. Abbreviations:
COMPASS-31, Composite Autonomic Symptom Score–31; BMI, body mass index.

When stratified by autonomic symptom burden, participants with COMPASS-31 ≥20 more frequently reported migraine (*P*=0.043), irritable bowel syndrome (*P*=0.036), hypotension (*P*=0.014), asthma (*P*=0.014), tinnitus (*P*=0.016), and generalized joint hypermobility (*P*=0.032), but not FMD (*P*=0.656). Diagnosed anxiety (*P*=0.048) and depression (*P*=0.028) were also more common in this group. Among traditional risk factors, current or former smoking differed by autonomic severity (*P*=0.011), whereas hypertension, dyslipidemia, alcohol use, and BMI ≥30 kg/m² did not (**Table 4**).

### Health-related quality of life, autonomic and fatigue severity symptoms

Moderate reductions in health-related quality of life were observed overall, with median scores of 71.0 (IQR 50.0–80.0) for EQ-VAS and 0.924 (IQR 0.848–0.956) for EQ-5D-5L utility (**Table 2**). Utility values ranged from −0.133 to 1.000, indicating substantial heterogeneity in health status. A ceiling effect was evident, with 13.7% of participants reporting the full health state (11111). Domain-level analysis demonstrated that health impairments were most frequently reported in pain/discomfort (80.2%) and anxiety/depression (63.9%), followed by limitations in usual activities (42.3%). Problems with mobility (24.2%) and self-care (6.2%) were less common. Fatigue burden was substantial, with a mean FSS score of 39.0±15.5 and 56.4% met criteria for severe fatigue (FSS ≥36).

Participants with COMPASS-31 ≥20 reported significantly poorer quality of life and greater fatigue. EQ-5D utility (0.904 [0.793–0.956] vs 0.924 [0.890–0.968], *P* <0.001) (**Figure 1A**), EQ-VAS (65.5 [50.0–75.0] vs 76.0 [62.0–82.0], *P* <0.001) (**Figure 1B**), and fatigue severity (45.5 [31.0–54.8] vs 33.0 [21.0–47.0], *P* <0.001) were all worse in the group with moderate-to-severe autonomic symptom burden. Mobility, self-care, and pain/discomfort impairment were also more frequent in this group (all *P* ≤0.029), whereas anxiety/depression did not differ according to autonomic symptom burden (*P*=0.187) (**Figure 2**). Greater autonomic symptom burden was moderately associated with lower EQ-5D utility (*r*ₛ=−0.337, *P* <0.001) (**Figure 3**) and independently predicted reduced health-related utility in univariable regression (*β*=−0.372, *P* <0.001; *R*²=0.139).

**Figure 1.**
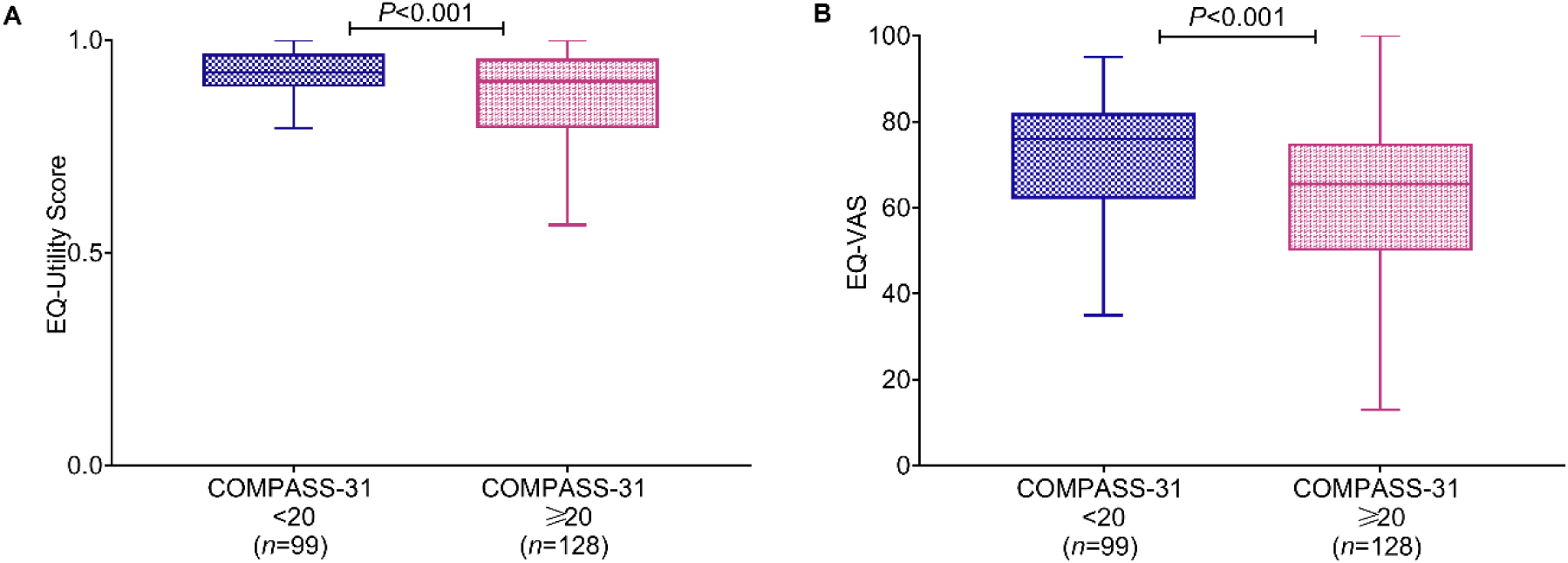
Health-related quality of life by autonomic symptom burden. Box-and-whisker plots showing EuroQol 5-Dimension 5-Level utility and EuroQol visual analog scale scores in participants with Composite Autonomic Symptom Score-31 <20 versus ≥20. Boxes represent the interquartile range, the horizontal line indicates the median.

**Figure 2.**
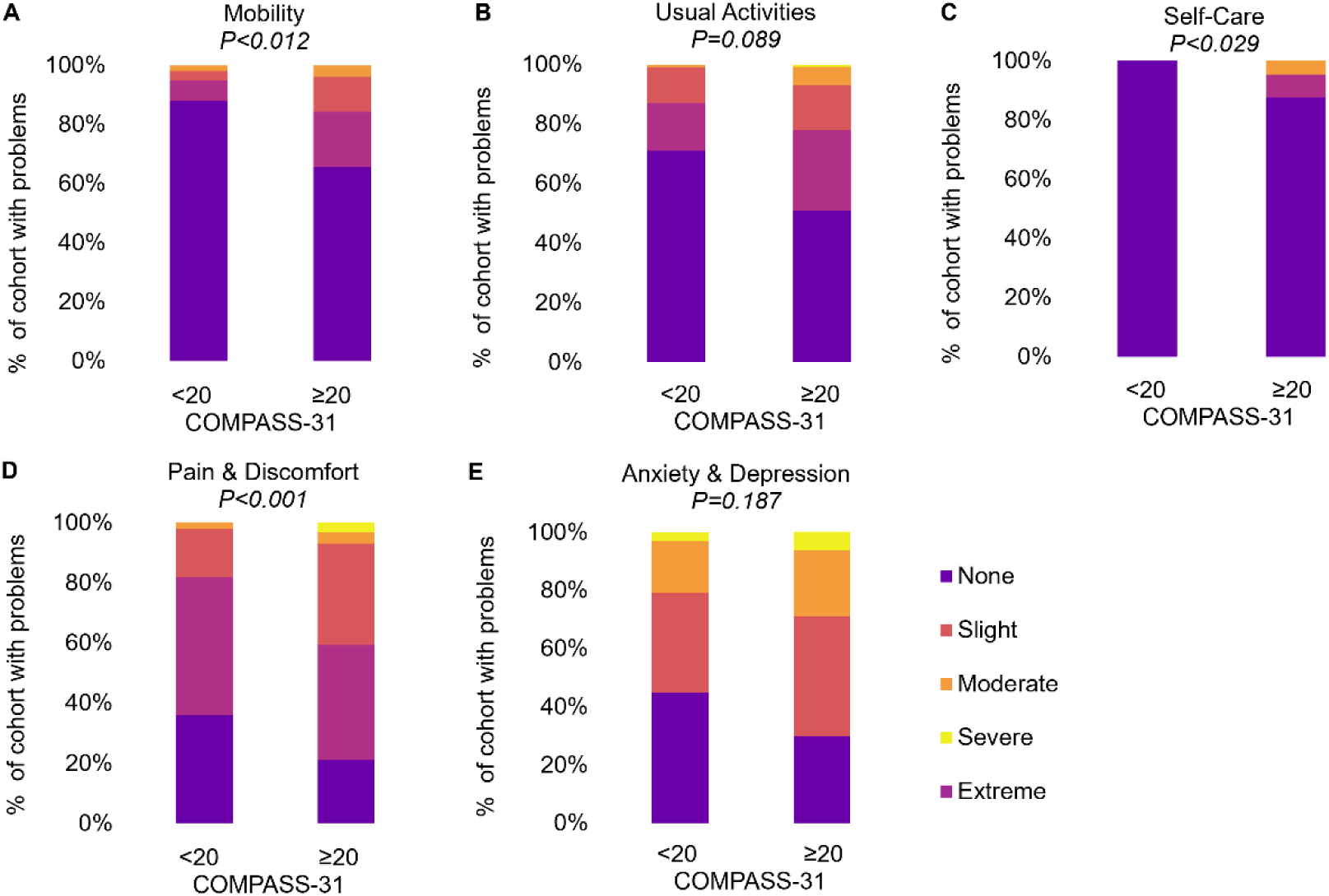
EuroQol-5 Dimension-5L domains by autonomic symptom burden. Stacked bar graphs illustrating the proportion of participants reporting impairment (levels 3–5) across EQ-5D-5L dimensions according to autonomic symptom burden (COMPASS-31 <20 vs ≥20). *n*=99 for COMPASS-31 < 20 and *n*=128 for ≥20. *P* values were derived from two-sided Pearson chi-square tests. Abbreviations: COMPASS-31, Composite Autonomic Symptom Score–31; EQ-5D-5L, EuroQol five-dimension five-level instrument.

**Figure 3.**
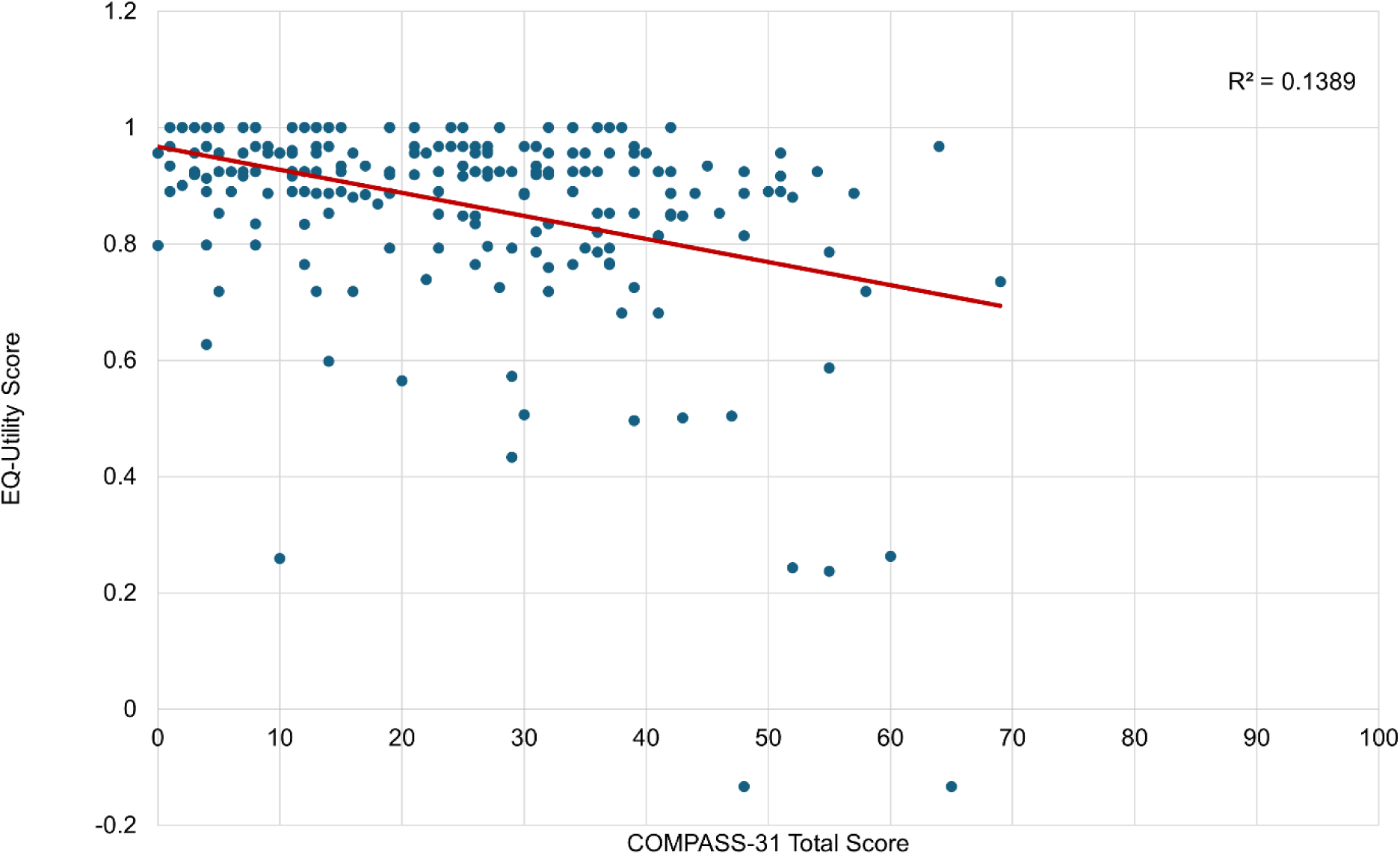
Association between autonomic symptom burden and health-related quality of life. Scatterplot demonstrates the association between COMPASS-31 total score and EQ-5D utility in 227 participants. Correlation was assessed using Spearman’s rho due to non-normal distribution of variables (*r*ₛ=−0.337, *P*<0.001). The superimposed line represents the linear regression model, which accounted for 13.9% of the variance in EQ-5D utility (*R²*=0.139).

In hierarchical multivariable regression analysis (**Supplementary Table 3**), demographic variables explained 16.1% of the variance in EQ-5D utility (adjusted *R*²=0.139). Addition of SCAD-specific variables did not improve model fit (Δ*R*²=0.000, *P*=0.998). However, inclusion of biological/autonomic comorbidity variables significantly increased explained variance (Δ*R*²=0.130, *P* <0.001), and inclusion of COMPASS-31 total score provided a further significant improvement (Δ*R*²=0.022, *P*=0.010). Addition of CES-D non-somatic score accounted for the largest final increment in explained variance (Δ*R*²=0.090, *P* <0.001), with the full model explaining 40.4% of the variance in EQ-5D utility (adjusted *R*²=0.364). In the final model, inability to work due to illness/disability (β=−0.201, *P* <0.001), fibromyalgia (β=−0.220, *P* <0.001), greater autonomic symptom burden measured by COMPASS-31 (β=−0.150, *P*=0.029), and higher CES-D non-somatic score (β=−0.331, *P* <0.001) were independently associated with lower EQ-5D utility. In contrast, CHAPS total score was not associated with EQ-5D utility after full adjustment (*P*=0.117).

### Anxiety and depression

The median ASI-3 total score in the cohort was 16 (9–26) out of a possible 72. Domain scores were 7 (4–12) out of 24 for physical concerns, 3 (1–8) out of 24 for cognitive concerns, and 5 (2–10) out of 24 for social concerns. In terms of anxiety sensitivity, endorsement was predominantly concentrated within the physical concerns domain. Moderate-to-high agreement (“some,” “much,” or “very much”) was reported by 60.3% of participants for rapid heart rate and by 66.9% for chest pain indicating a heart attack. Similarly, 45.3% of participants endorsed moderate-to-high concern regarding palpitations. In contrast, cognitive concern items were predominantly endorsed at minimal levels. For example, 56.8% of participants endorsed “very little” fear of going crazy when unable to concentrate, 70.5% endorsed “very little” fear of losing control of thoughts, and 69.2% “very little” fear of mental incapacitation.

When stratified by autonomic symptom burden (COMPASS-31 <20 vs ≥20), responses to several ASI-3 physical concerns significantly differed between groups, including tachycardia, chest pain–related concern, and palpitations (all *P* ≤0.05), with greater endorsement of higher severity categories in the ≥20 group (**Figure 4**).

**Figure 4.**
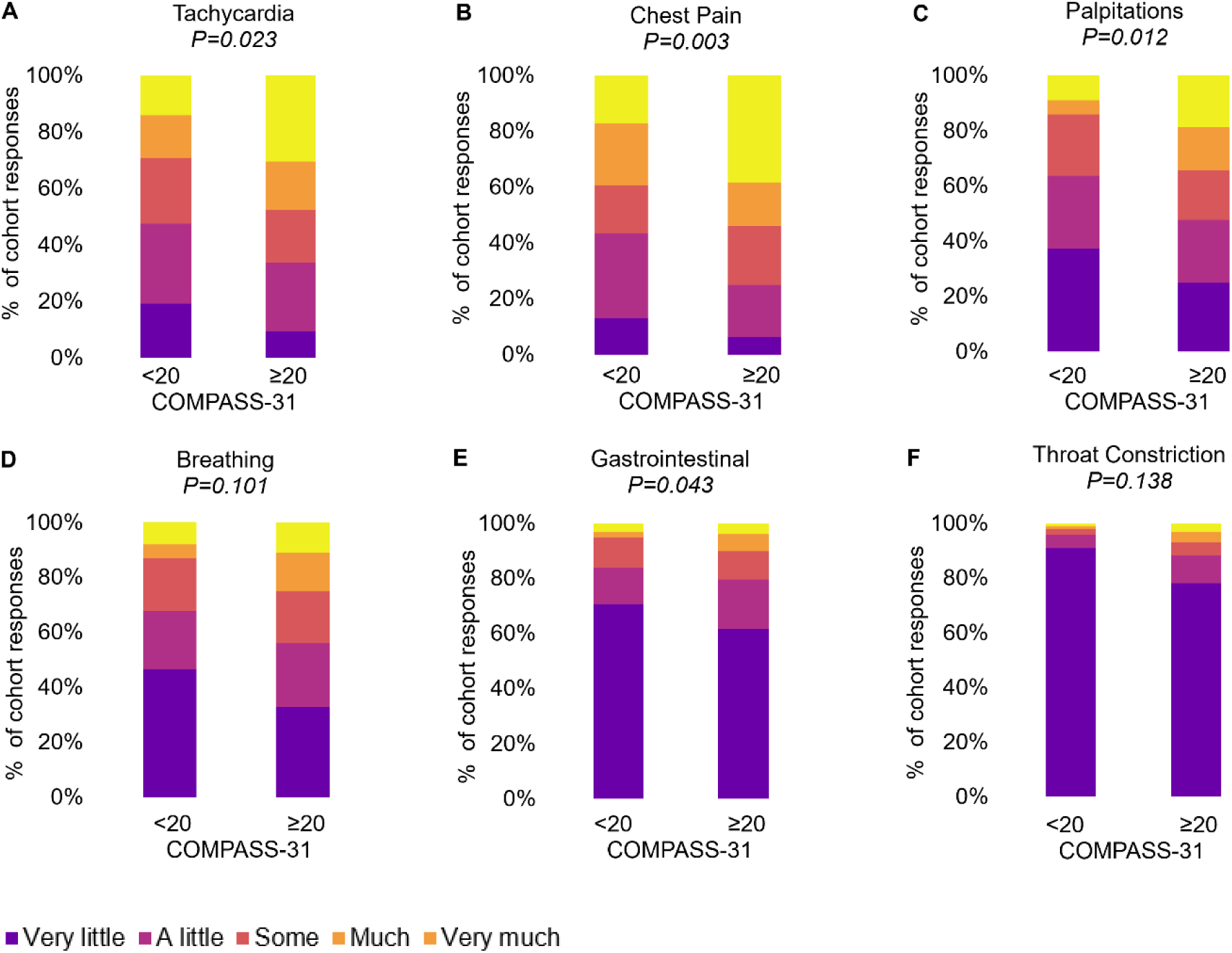
Distribution of ASI physical concerns item responses by autonomic symptom burden. Stacked bar charts illustrating response distributions for six Anxiety Sensitivity Index-3 (ASI-3) physical concerns items stratified by Composite Autonomic Symptom Score (COMPASS-31) total score (<20 vs ≥20). Bars represent the percentage of participants endorsing each response category (“very little” to “very much”). *P* values reflect Pearson *χ²* tests comparing distributions between groups.

Depressive symptom burden was also higher among participants with COMPASS-31 ≥20 (median 18 [9–27] vs 11 [5–17], *P* <0.001). This also applied to non-somatic CES-D scores, although the magnitude of difference was smaller than for total CES-D score. Item-level analysis showed that the clearest between-group differences were in somatic and psychomotor symptoms, particularly reduced appetite, trouble concentrating, fatigue/effort, restless sleep, and difficulty getting going (**Supplementary Table 4**). Reduced positive affect and some affective symptoms, including feeling depressed, lonely, and sad, were also more common in the higher autonomic symptom burden group, whereas several other mood and interpersonal items showed little difference. Using a CES-D ≥20 threshold, 31.7% of participants met the criteria for clinically significant depressive symptoms; this proportion decreased to 13.2% when somatic items were excluded.

In multivariable regression adjusted for depressive symptoms, COMPASS-31 was independently associated with ASI-3 total score (*β*=0.214, *P*<0.001) and domain-specific physical (*β*=0.232, *P*<0.001) and cognitive concerns (β=0.181, *P*=0.004), but not social concerns (*P*=0.060) **(Table 5**), with CES-D the stronger predictor across all domains. Depressive symptoms were significantly associated with all ASI-3 domains. In sensitivity analyses, combined cognitive and social ASI-3 domains showed a weaker association with COMPASS-31 (*β*=0.165, *P*=0.008), whereas depressive symptoms remained the dominant predictor (*β*=0.441).

**Table 5.** Multivariable linear regression models examining associations between autonomic symptom burden and anxiety sensitivity domains (*n*=227)

| **Outcome** | **Predictor** | **Standardized $\beta$** | **$P$ value** | **Adjusted $R^2$** |
| --- | --- | --- | --- | --- |
| ASI Physical | COMPASS-31 | 0.232 | **< 0.001** | 0.196 |
|  | CES-D | 0.308 | **< 0.001** |  |
| ASI Cognitive | COMPASS-31 | 0.181 | **0.004** | 0.255 |
|  | CES-D | 0.415 | **<0.001** |  |
| ASI Social | COMPASS-31 | 0.122 | 0.060 | 0.197 |
|  | CES-D | 0.392 | **< 0.001** |  |

All models are adjusted for depressive symptoms (CES-D total score). Standardized $\beta$ coefficients are reported. Abbreviations: ASI, Anxiety Sensitivity Index–3; CES-D, Center for Epidemiologic Studies Depression Scale; COMPASS-31, Composite Autonomic Symptom Score–31.

## Discussion

This is the first study to systematically quantify autonomic symptom burden in survivors of SCAD. Our findings are as follows: i) More than half of individuals with SCAD demonstrated at least moderate autonomic symptom burden, with greatest impairment in orthostatic intolerance and gastrointestinal domains. ii) Autonomic symptom burden was independently associated with reduced health-related quality of life and severe fatigue, with impairment concentrated in physical rather than affective domains. iii) Anxiety sensitivity was predominantly characterized by cardiac-focused physical concerns, with no independent association between autonomic burden and social anxiety features. iv) Contemporary autonomic symptom burden was independently predicted by antecedent autonomic features, particularly orthostatic symptoms in adolescence and young adulthood.

### Cohort characteristics in context

The demographic profile of our cohort was consistent with the recognized SCAD phenotype. Participants were predominantly women presenting in midlife, with age at first event centered in the early 50s, in keeping with contemporary Canadian and Australian–New Zealand registry data.^27, 28^ The proportions of women and White participants were higher in this study, probably reflecting recruitment methods and English-language survey participation rather than true epidemiologic differences. Expectedly, traditional cardiovascular risk factors were infrequent while conditions linked to vascular lability were more prominent. In particular, migraine prevalence was consistent with established SCAD registries,^27, 28^ although FMD was reported less often than in CanSCAD,^28^ likely due to differences in extra-coronary screening. Recurrent SCAD was experienced in over a fifth of our participants, who were a median of three years from their index event at the time of survey.

Examination of event timing and reported precipitants provides additional context for SCAD presentation. Precipitating stressors were notable for the predominance of emotional triggers. Intense emotional stress was reported far more often than intense physical stress, consistent with prior registry findings, including from CanSCAD.^28^ Emotional precipitants mostly involved relational conflict, occupational strain, bereavement, or serious family illness. Together with qualitative literature describing SCAD as abrupt and often occurring during acute life stress, these findings support emotional stressors as a reproducible clinical feature of SCAD presentation rather than solely retrospective attribution.^27–30^

### Autonomic dysregulation and vascular susceptibility in SCAD

In the present study, autonomic symptom burden was quantified using COMPASS-31, a validated multidomain instrument widely used to assess clinically meaningful autonomic symptoms across neurologic and cardiovascular conditions.^15, 31, 32,33^ Within this framework, the autonomic signal in SCAD survivors was substantial. More than half reported at least moderate autonomic symptom burden, and one in six had severe burden. Cardiovascular and gastrointestinal symptoms predominated with additional vasomotor and secretory features, suggesting broader neurovascular involvement rather than isolated positional intolerance. These rates exceed those reported in healthy populations and differ from atherosclerotic coronary artery disease, where autonomic abnormalities more often reflect impaired heart rate variability and baroreflex function over multidomain systemic symptom burden.^11, 34, 35^ Another important finding was that antecedent autonomic features in adolescence and young adulthood were independently associated with current autonomic symptom burden. The CHAPS remained the strongest independent predictor after adjustment for demographic factors, cardiovascular risk profile, SCAD characteristics, and comorbidities, with orthostatic dizziness showing the strongest association. This prompts speculation that there may be pre-existing autonomic vulnerability in patients who experience SCAD, rather than autonomic dysfunction being purely reactive to the acute SCAD event.

This interpretation fits the broader clinical picture of SCAD, which clusters with disorders marked by vasomotor instability and dysregulated neurovascular control. Migraine is overrepresented in SCAD populations,^27, 28, 36^ and functional gastrointestinal disturbance, a recognized manifestation of enteric autonomic dysregulation,^37, 38^ was also common in our cohort. The co-occurrence of migraine, gastrointestinal disturbance, hypermobility, and multidomain autonomic burden supports a shared regulatory substrate rather than isolated organ-specific pathology.

Autonomic dysregulation may be particularly relevant in the setting of structural arterial susceptibility. Enrichment of rare variants in vascular connective tissue genes among high-risk SCAD phenotypes supports intrinsic arterial fragility in a subset of patients.^5, 39^ In this context, even modest disturbances in sympathetic–parasympathetic balance, blood pressure variability, or episodic vasomotor instability may assume disproportionate pathophysiologic significance. The interaction between dysregulated autonomic control and a structurally susceptible arterial wall offers a biologically plausible model integrating regulatory and structural vulnerability. However, autonomic symptom burden was not independently associated with SCAD recurrence. This suggests that while dysautonomic traits may contribute to initial vulnerability to SCAD or persistent symptoms post-MI, recurrence may be driven by other factors.

### Anxiety, depression and autonomic symptom overlap

Anxiety and depressive symptoms are frequently reported after SCAD, and prior studies suggest psychosocial distress may be more common than after atherosclerotic MI.^40^ However, elevated affective scores require careful interpretation. Large angiographic registries report relatively modest rates of clinician-documented anxiety and depression, whereas our cohort showed higher rates of practitioner-diagnosed affective disorders.^27, 28^ This probably reflects differences in ascertainment, survivorship duration, and measurement rather than true epidemiologic variation. Importantly, lifetime clinical diagnosis and current symptom burden are not equivalent. In this cohort, moderate anxiety/depression scores likely reflected a combination of recurrence-related uncertainty and ongoing symptom burden rather than pervasive affective disorder.

Anxiety sensitivity scores were driven mainly by cardiac-focused physical concerns, including tachycardia, chest discomfort, and fear of cardiac catastrophe, rather than cognitive fears of losing control. Social concern items attenuated after adjustment for depressive symptoms, supporting heightened vigilance to recurrent physiological sensations rather than a primary cognitive anxiety phenotype. A similar pattern was seen for depressive symptoms. Although nearly one third of the cohort met criteria for clinically significant depressive symptoms using a conservative medical threshold, removal of somatic items substantially reduced prevalence, indicating considerable symptom overlap. Fatigue, sleep disturbance, and cognitive difficulty were most common, whereas positive affect remained relatively preserved, suggesting symptom-driven impairment rather than pervasive anhedonia or sustained low mood. These findings align with autonomic and orthostatic disorders, in which hemodynamic instability and cerebral hypoperfusion can contribute to fatigue and cognitive symptoms independent of primary mood disorder.^18, 26, 41^ In this context, elevated anxiety and depression scores may partly reflect interoceptive vigilance and lived physiologic instability.^19^ Careful distinction between expression of physiologic symptoms and primary affective illness is therefore essential in patients with SCAD, particularly in the setting of autonomic vulnerability.

### Health-related quality of life

Health-related quality of life in our cohort was largely preserved at a global level. Using the contemporary Australian Norman EQ-5D-5L value set, mean utility was 0.924, with a negatively skewed distribution indicating clustering toward preserved health states. Even so, moderate impairment in self-rated health was evident, and fatigue burden was substantial, with more than half of participants meeting criteria for severe fatigue. These findings are consistent with longitudinal data from the VIRGO registry, in which SCAD survivors demonstrated similar or higher EQ-5D scores than other MI cohorts over time.^36^ By contrast, a recent Australian survey reported lower utility estimates, likely reflecting use of the EQ-5D-3L and different valuation algorithms, which can materially affect utility estimates in cohorts with predominantly mild-to-moderate impairment.^24, 42, 43^

Autonomic symptom burden was an important correlate of reduced health-related quality of life. Participants with moderate autonomic dysfunction reported lower health utility, poorer self-rated health, and greater fatigue, with impairment concentrated in physical domains, such as mobility and pain rather than anxiety or depression. In hierarchical models, autonomic symptom burden remained independently associated with lower health utility after adjustment for demographic and clinical factors, although depressive symptoms showed the strongest independent association. Together, these findings suggest that residual symptom burden after SCAD may be influenced in part by autonomic dysfunction, particularly in relation to fatigue and physical limitation.

### Future considerations

The current findings suggest that autonomic symptom burden in SCAD reflects a broader systemic phenotype rather than a nonspecific post-event effect. Although causality cannot be inferred from our cross-sectional data, they do provide a rationale for prospective mechanistic studies incorporating objective autonomic and vascular phenotyping in this poorly understood condition.^41^ Clinically, assessment of autonomic symptoms could also help identify SCAD patients who may benefit from targeted management of orthostatic intolerance and related symptoms to improve quality of life, fatigue and physical limitation during their short and longer term recovery.

### Limitations

Our study has several limitations. Its cross-sectional design precludes inference about temporality or causality, and historical autonomic features were reported retrospectively, introducing potential recall bias. Although validated instruments were used to assess autonomic and psychological symptoms, fatigue, and health-related quality of life, all measures were self-reported and objective autonomic testing was not performed. These instruments capture perceived rather than physiological burden. Medication usage was not collected, and autonomic-active therapies may have influenced orthostatic symptoms, although this would not explain the association between antecedent and current autonomic features. SCAD diagnosis and comorbidities were also self-reported as clinician-diagnosed and were not independently verified. Recruitment through clinician networks and consumer organizations may have influenced participant characteristics, and the cohort was predominantly female and White, limiting generalizability. These findings should therefore be considered hypothesis-generating and require confirmation in prospective studies incorporating objective autonomic assessment.

### Conclusion

Autonomic symptom burden is common and clinically meaningful in survivors of SCAD, even years after the index event. Autonomic dysregulation may represent a pre-existing vulnerability in at least a subset, rather than solely a reactive consequence of the event. Although causality cannot be inferred, the convergence of historical predisposition, contemporary symptom burden, and physical quality-of-life impairment suggests that autonomic dysfunction may be relevant to the clinical phenotype of SCAD and warrants further prospective investigation.

## Acknowledgements

An AI-based language model was used to support grammar, readability, and formatting of tables and text. All intellectual content, data analysis, and interpretation were generated and verified by the authors. We gratefully acknowledge the individuals with spontaneous coronary artery dissection whose participation made this study possible.

